# NF1-specific growth charts for head circumference over the first three years of life

**DOI:** 10.1101/2025.06.23.25328558

**Authors:** Ayan S. Mandal, Russell T. Shinohara, Benjamin Jung, Margaret Gardner, Habib E. Akouri, Benjamin E. Yerys, Whitney Guthrie, Kelly M. Janke, John D. Herrington, Matthew C. Hocking, Gareth Ball, Jonathan M. Payne, Kathryn N. North, Nils Muhlert, Shruti Garg, Jakob Seidlitz, Michael J. Fisher, Aaron Alexander-Bloch

**Author notes:** Please address correspondence either to Ayan Mandal or to Aaron Alexander-Bloch: Richards Medical Research Laboratories, 3700 Hamilton Walk, Philadelphia, PA 19104. **Disclosures:** Drs. Seidlitz and Alexander-Bloch hold shares in and Drs. Seidlitz is a director of Centile Bioscience.

## Abstract

**Background and objectives:** Macrocephaly is among the most common findings in neurofibromatosis type 1 (NF1) and may be associated with other clinical manifestations of the genetic syndrome. NF1-specific growth charts that account for expected macrocephaly may increase sensitivity to detect atypical growth. We aimed to produce NF1-specific growth charts of head circumference for the age range of 0 to 3 years and to assess their potential clinical impact.

**Methods:** Using electronic health records from the Children’s Hospital of Philadelphia, we collected head circumference measurements from children with NF1 and a community control cohort seen at scheduled well-child visits. We compared head circumference normed using Center for Disease Control (CDC) growth charts between these groups over time. We constructed NF1-specific growth charts using two independent methods. Finally, we used mixed-effects models to relate the resulting centile scores with developmental delay assessed with the Survey of Well-being of Young Children.

**Results:** Our dataset contained 2180 observations from 305 individuals (167 male) with NF1, and 104,750 observations from 16,742 individuals (8809 male) in the community control cohort, all aged 0 to 3 years old. Head circumference was significantly elevated in NF1 throughout the age range (*P*_adjusted_ <0.05), but the effect sizes varied nonlinearly with age, starting moderate at 1 month (*d* = 0.56), then small at four months (*d* = 0.28), moderate again at 15 months (*d* = 0.58), and finally large at 28 months (*d* = 0.8). NF1-specific growth curves demonstrated slower rate-of-growth for head circumference in the first two months of life yet more sustained growth over time. Although none of the children with NF1 met the standard for microcephaly according to CDC charts, smaller head circumference benchmarked against NF1-specific charts was correlated with developmental delay (standardized beta = 0.24; *P* = 0.013).

**Discussion:** We present the first NF1-specific growth charts for head circumference covering ages 0 to 3 years. Macrocephaly in NF1 becomes more exaggerated over time as rate-of-growth is sustained compared to controls. Smaller head size relative to NF1 growth expectations is not captured by CDC charts yet nevertheless relates to developmental delay, suggesting that NF1-specific charts may increase sensitivity to clinically concerning patterns of growth in children with NF1.

## Introduction

Neurofibromatosis type 1 (NF1) is a genetic syndrome affecting roughly 1:3000 births^1^ with a wide range of clinical manifestations, including plexiform neurofibromas, low grade gliomas, cognitive issues, and macrocephaly.^2,3^ These oncologic and non-oncologic manifestations are thought to result from overactivation of the RAS pathways due to loss of function of the *NF1* gene.^3,4^ The oncologic manifestations of NF1 are explained by the classic two-hit hypothesis for tumor suppression syndromes, as loss of the second *NF1* allele is sufficient to trigger tumor formation.^4,5^ Haploinsufficiency of the *NF1* gene is hypothesized to cause the non-oncologic manifestations of NF1, including macrocephaly, megalencephaly, and cognitive symptoms among many others, yet the exact mechanisms remain poorly understood.^6,7^

Macrocephaly, defined as a head circumference two standard deviations above the population reference average for age and sex, is one of the most common neurological manifestations of NF1, affecting an estimated 37.5% of patients.^8^ Head circumference is strongly coupled with total brain volume in the early age range among both healthy and clinical populations.^9,10^ Macrocephaly in NF1 is primarily attributed to megalencephaly, with meta-analytic findings demonstrating increased global brain volumes across both grey and white matter, even in the absence of tumors.^11,12^ In turn, larger white matter and subcortical gray matter volumes in NF1 have been associated with deficits in social cognition and executive functioning.^13^ However, little is known regarding the timing of deviations in brain growth in NF1 compared to typical development.

Typical head growth is accelerated in early infancy over the first few months of life before stabilizing and eventually plateauing around two years of age.^14^ Early brain overgrowth within the first two years of life is a hypothesized (albeit controversial^15^) mechanism of divergent brain development in autism spectrum disorder (ASD),^16,17^ which co-occurs with NF1 in 10-30% of cases.^18,19^ It is unclear whether macrocephaly in NF1 is the result of more rapid head growth in the early months of life, or a longer, more sustained period of elevated head growth compared to what is seen in typical development.

Furthermore, head circumference growth charts specific to NF1 could be a helpful clinical tool as standard growth charts (e.g., those provided by the Center for Disease Control (CDC) or World Health Organization (WHO)) may not be appropriate for these patients.^20^ For example, an extremely high percentile for head circumference based on a standard growth chart applied to an NF1 patient may be attributed to their genetic condition, leaving other causes of abnormally elevated head circumference (e.g., obstructive hydrocephalus) uninvestigated. Conversely, an NF1 patient with a normal percentile for head circumference based on a standard growth chart may actually be microcephalic relative to their NF1 peers, warranting further clinical evaluation. While NF1 head circumference growth charts have been previously reported covering ages ranging from 2 to 19 years,^21,22^ no publicly available charts to our knowledge currently cover the first two years of life, the period of time before closure of the fontanelles when head circumference is most closely monitored clinically.

Here, we present the first publicly available head circumference growth charts for NF1 patients aged 0 to 3 years. We describe two independent approaches to defining NF1 head circumference growth charts that provide complementary insights on NF1 head development. Finally, we relate head circumference benchmarked upon NF1 growth charts to measures of developmental milestone completion to validate the clinical utility of NF1-specific growth charts in uncovering otherwise obscured growth abnormalities.

## Methods

### Cohort Selection

Using electronic health records from the Children’s Hospital of Philadelphia (CHOP), we accessed 110,927 head circumference measurements from 17,307 patients aged 0 to 3 years old seen at either a well-child visit (for routine pediatric follow up) or who were NF1 patients seen by the CHOP Neurofibromatosis Multidisciplinary Program. In accordance with the American Academy of Pediatrics recommendations, well child visits at CHOP are scheduled at the first week of life, 1 month old, 2 months old, 4 months old, 6 months old, 9 months old, 1 year old, 15 months old, 18 months old, 2 years old, 30 months old, and 3 years old. Head circumference data were taken from a relational database enriched for patients who had received an MRI scan at CHOP.^23^ Head circumference observations were obtained between the years 2003 and 2025.

Using each participant’s gestational age at birth and age at the time of each observation, we calculated their post-conceptional age (e.g. a 30 day old child born at 40 weeks had a post-conceptional age of 310 days). Gestational age at birth was assumed to be 40 weeks for the 41 NF1 participants and 1719 clinical controls for whom it was not available. We excluded individuals born at less than 26 weeks of gestation, with birth weight less than 1 kg, and/or whose post-conceptional age was less than 252 days (corresponding to 36 weeks). This resulted in a dataset of 107,999 observations from 16,779 individuals.

Next, we sought to remove implausible head circumference measurements that likely reflected data entry errors. To do this, we used an outlier detection approach applied previously by our group^10^ to calculate modified Z-scores for each head circumference observation from the whole pooled study population. This involved first fitting a b-spline with four degrees of freedom to the head circumference data using post-conceptional age and sex as predictors. We used the residuals of this model as the numerator for the modified Z-score. The denominator included the median of the absolute deviations (MAD, or median of the absolute values of the residuals) multiplied by a scaling factor set to 1.4826 assuming normally distributed data. We visualized the modified Z-scores on a histogram (Supplementary Figure 1) and excluded observations where the absolute value of the score exceeded 5. This resulted in a dataset of 107,412 observations from 16,746 individuals.

We also sought to remove within-subject outlier head circumference measurements that were also thought to be implausible and likely reflecting data entry error. To that end, we calculated for each observation the difference between the modified Z score and the median modified Z score across observations from the same participant. We plotted these values on a histogram (Supplementary Figure 2) and excluded observations where the absolute value of the difference exceeded 2. The resulting number of observations in the dataset was 106,970 from 16,744 individuals. Finally, we removed data from four individuals where sex was documented as neither male nor female, resulting in a final dataset of 106,950 observations from 16,742 individuals (Figure 1; Table 1).

**Figure 1.**
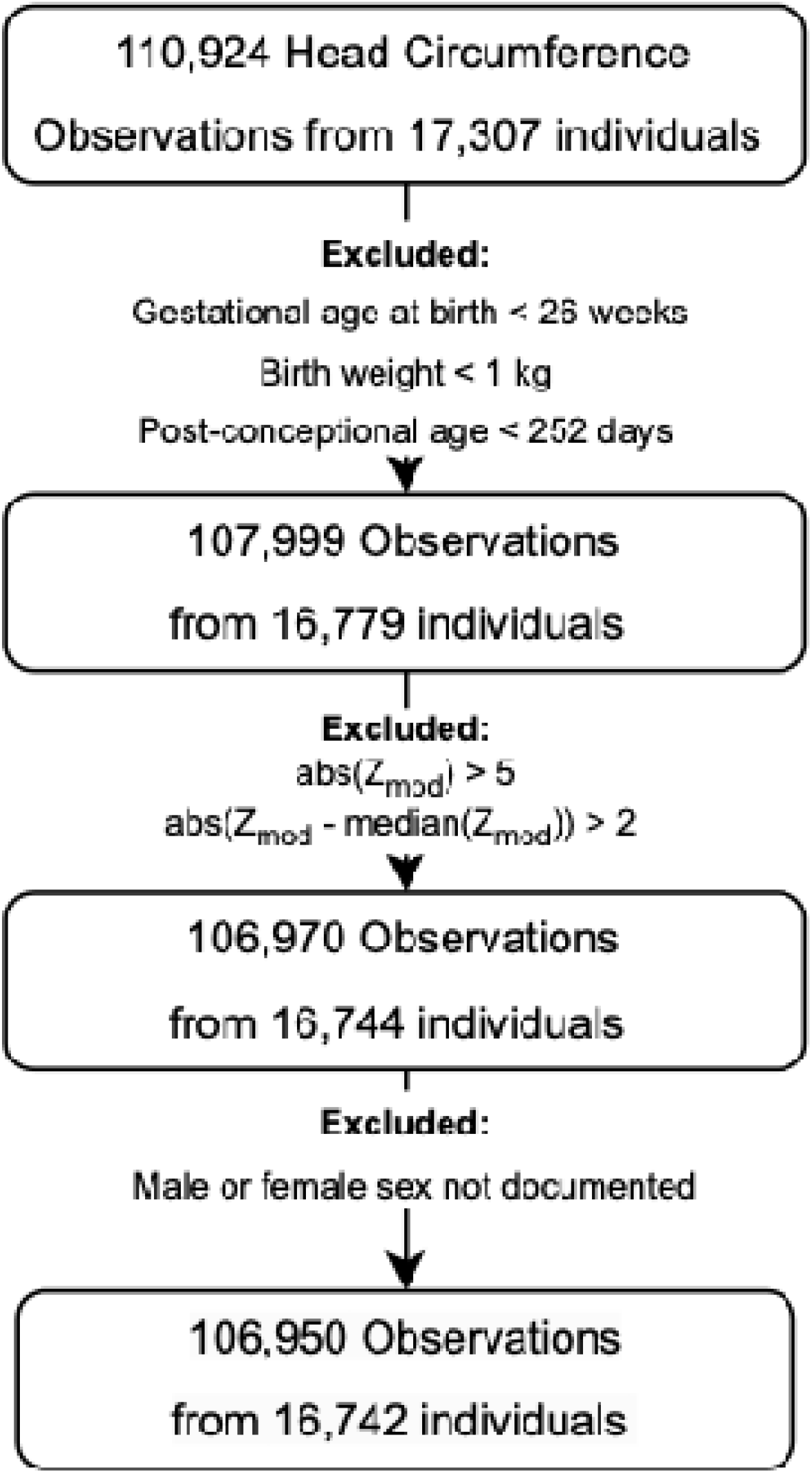
Flow chart summarizing cohort selection for Well Child and NF1 head circumference data.

**Table 1.**
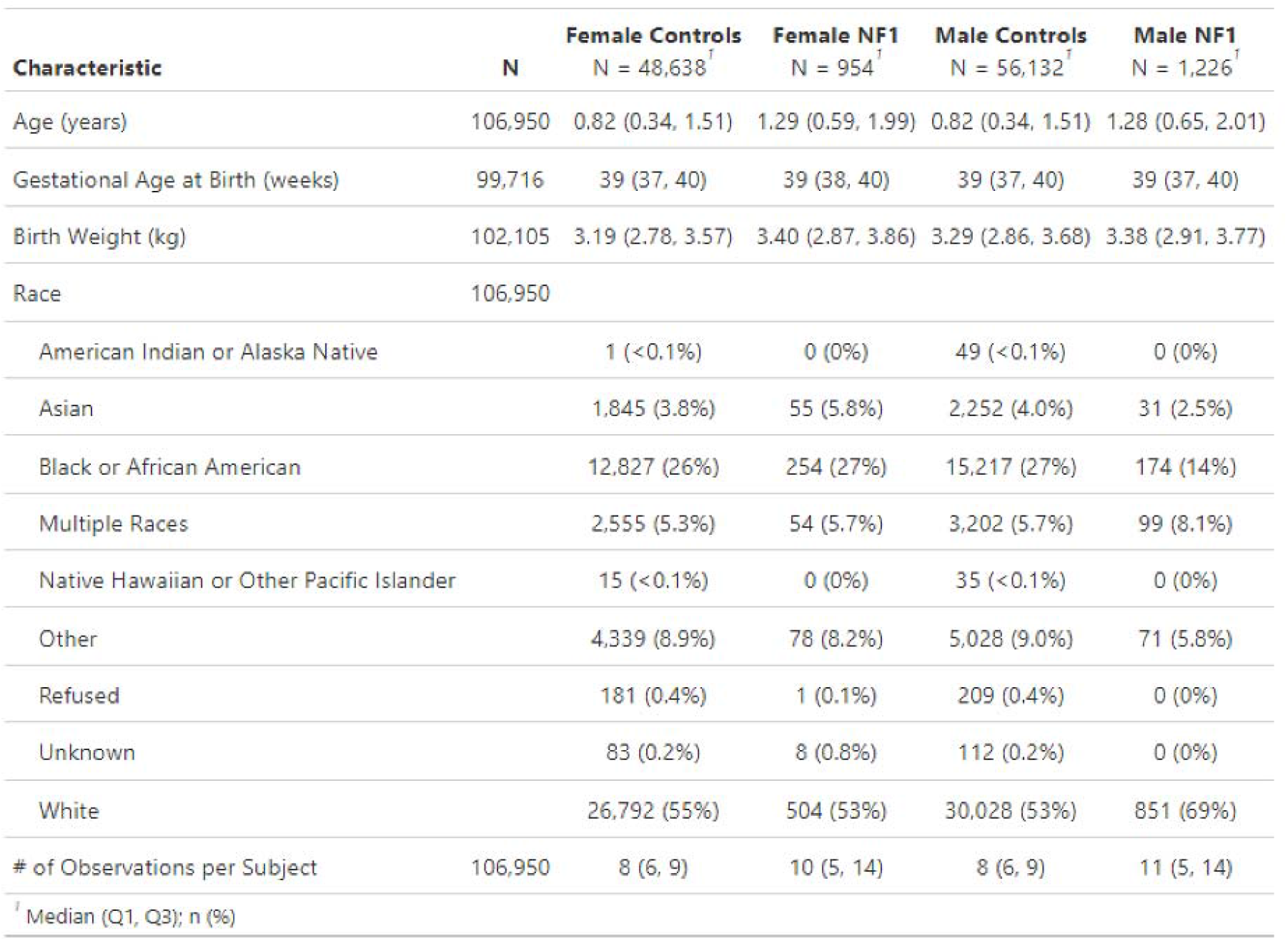
Demographics for Head Circumference Observations in each cohort. These data describe demographic information corresponding to the number of head circumference observations within each cohort, for which there were multiple observations per participant. In the Female Controls, Female NF1, Male Controls, and Male NF1 cohorts, there were 7628, 138, 8809, and 167 unique participants respectively.

| Characteristic | N | Female Controls<br>N = 48,638<sup>†</sup> | Female NF1<br>N = 954<sup>†</sup> | Male Controls<br>N = 56,132<sup>†</sup> | Male NF1<br>N = 1,226<sup>†</sup> |
| --- | --- | --- | --- | --- | --- |
| Age (years) | 106,950 | 0.82 (0.34, 1.51) | 1.29 (0.59, 1.99) | 0.82 (0.34, 1.51) | 1.28 (0.65, 2.01) |
| Gestational Age at Birth (weeks) | 99,716 | 39 (37, 40) | 39 (38, 40) | 39 (37, 40) | 39 (37, 40) |
| Birth Weight (kg) | 102,105 | 3.19 (2.78, 3.57) | 3.40 (2.87, 3.86) | 3.29 (2.86, 3.68) | 3.38 (2.91, 3.77) |
| Race | 106,950 |  |  |  |  |
| American Indian or Alaska Native |  | 1 (<0.1%) | 0 (0%) | 49 (<0.1%) | 0 (0%) |
| Asian |  | 1,845 (3.8%) | 55 (5.8%) | 2,252 (4.0%) | 31 (2.5%) |
| Black or African American |  | 12,827 (26%) | 254 (27%) | 15,217 (27%) | 174 (14%) |
| Multiple Races |  | 2,555 (5.3%) | 54 (5.7%) | 3,202 (5.7%) | 99 (8.1%) |
| Native Hawaiian or Other Pacific Islander |  | 15 (<0.1%) | 0 (0%) | 35 (<0.1%) | 0 (0%) |
| Other |  | 4,339 (8.9%) | 78 (8.2%) | 5,028 (9.0%) | 71 (5.8%) |
| Refused |  | 181 (0.4%) | 1 (0.1%) | 209 (0.4%) | 0 (0%) |
| Unknown |  | 83 (0.2%) | 8 (0.8%) | 112 (0.2%) | 0 (0%) |
| White |  | 26,792 (55%) | 504 (53%) | 30,028 (53%) | 851 (69%) |
| # of Observations per Subject | 106,950 | 8 (6, 9) | 10 (5, 14) | 8 (6, 9) | 11 (5, 14) |

<sup>†</sup> Median (Q1, Q3); n (%)

### Comparing Head Circumference Z Scores between NF1 and Well Child Cohort

We used the LMS method from the Center for Disease Control (CDC) growth charts on head circumference to compute Z scores for the well-child and NF1 cohorts.^24^ We used the CDC charts because they are used in clinical practice at CHOP as well as many other pediatric medical centers. Because the input to LMS for the CDC charts is age represented in months, we transformed post-conceptional age by subtracting 280 days, dividing by 30.437, rounding down to an integer, and adding 0.5. As such, the age of a 7-week-old child born at term would be represented as 1.5 months on the CDC charts, as would all children with age between 1.0 and 1.99 months of age corrected for gestational age at birth.

Next, we compared CDC head circumference Z scores between the NF1 and well-child cohort at the timing of each well-child visit, where the head circumference data are most dense. Head circumference observations were grouped into bins based on whether that observation was 10 days plus or minus the age corresponding to the well-child visit timepoint. If multiple head circumference observations from the same individual appeared within the same bin, the Z score of the observation closest to the well-child visit timepoint was chosen. Unpaired two-sample T tests were conducted between both groups at each age bin, with multiple comparisons corrected using Holm-Bonferroni correction. Given the large sample sizes, Cohen’s *d* was also calculated for each comparison to quantify effect sizes which are more interpretable within this context.^25^

### Construction of NF1-Specific Head Circumference Growth Charts using LMSz

Next, we implemented a recently described method called “LMSz” to construct NF1-specific head circumference growth charts.^26^ Following the procedure described by Low and colleagues, we fit generalized additive models for location, scale, and space (GAMLSS)^27^ with the normal distribution family to CDC head circumference Z-scores for the NF1 observations using age and sex as covariates. Head circumference observations were inverse weighted by the total number of observations per subject. We iterated through a set of candidate models for the mu and sigma formulas within GAMLSS and computed Akaike Information Criteria (AIC) and Bayesian Information Criteria (BIC) for each (Supplementary Table 1). For the final NF1-specific charts, we chose the model that minimized BIC as this model was simpler than the one that minimized AIC. The resulting model included age as a linear term for mu and only the intercept term for sigma. Using this GAMLSS model, we computed centile lines for the 3^rd^, 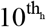, 50^th^, 90^th^, and 97^th^ percentile for CDC Head Circumference Z score, then transformed these centile lines back into head circumference measurements using the LMS method. We also examined rate-of-growth by computing the derivative of the 50^th^ percentile line of the NF1-specific and CDC growth charts respectively. GAMLSS model visualization throughout the study was performed using customized R functions available in gamlssTools (https://github.com/BGDlab/gamlssTools).

### Deriving CHOP Growth Charts for NF1 and Community Controls

While widely used in clinical practice, CDC growth charts have potential limitations. First, clinical head circumference data (even for well-child visits) may deviate systematically from CDC norms^15,28^. Second, for neonatal head circumferences in premature infants, CDC growth charts are suboptimal because they start at birth and are not corrected for gestational age. To address these limitations, we also computed separate growth charts using the NF1 and well-child visit data, henceforth referred to as “CHOP growth charts” (which, in contrast to the LMSz charts, only use data from the CHOP cohorts). Following the protocol described by the WHO,^29^ we fit natural splines for the mu and sigma moments in GAMLSS with degrees of freedom ranging from 5 to 15, and 2 to 10 respectively. Tau and nu were fit with an intercept term, and the Box-Cox Power Exponential (BCPE) distribution was used. In addition, we also computed models where penalized b-splines were used for mu and sigma. Age was represented as the logarithm (base of 10) of post-conceptional age measured in days. Head circumference observations were inverse weighted by the total number of observations per subject.

We computed separate GAMLSS models for males in the well-child cohort (Male Controls), females in the well-child cohort (Female Controls), males in the NF1 cohort (Male NF1), and females in the NF1 cohort (Female NF1). For each of these cohorts, we identified the models that produced the lowest AIC and BIC as well as the result of the penalized b-spline model and chose one of the three based on the fit of the worm plot and smoothness of the curves. If these criteria were equal across multiple options, we chose the simplest of the remaining models. Based on this process, a model with five degrees of freedom for mu and two degrees of freedom for sigma was selected for each cohort (Supplementary Figures 3-6). We also visualized quantile-quantile (Q-Q) plots for each selected model (Supplementary Figure 7). We compared Z scores from the LMSz and CHOP growth charts (Supplementary Figure 8). Finally, we applied both the LMSz and CHOP NF1-specific growth charts to an independent NF1 cohort recruited in Australia to test the generalizability of the charts (see Supplementary Materials).

### Relating Head Circumference Benchmarked Against NF1 Growth Charts to Developmental Delay

We examined whether head circumference benchmarked against NF1 growth charts was more sensitive to developmental delay in NF1 patients compared to standard growth charts. First, we aligned the NF1 head circumference data to Survey of Well Being of Young Children (SWYC) surveys conducted on the same days, which assess developmental milestones (specifically gross motor, fine motor, language, cognitive, and social development) at pediatric primary care visits.^30^ The SWYC data were converted into a developmental quotient (DQ) using a previously-described normative model to quantify the extent to which a child has met expected milestones for their age.^31^ The DQ is the estimated age of the child based on the number of expected milestones they have achieved divided by their chronological age (e.g. a 12 month old who has only met the milestones expected of an 8 month old has a DQ of 0.75). Using linear mixed effects models, we evaluated the relationship between DQ and head circumference Z-scores from the LMSz NF1 charts where patient ID was used as a random effect. We also plotted histograms of NF1 head circumference benchmarked against the CDC and LMSz charts respectively to evaluate prevalence of microcephaly (i.e. head circumference Z score < 2) from either approach. Finally, we assessed the relationship between DQ and head circumference Z scores derived from the CHOP NF1 charts as well.

### Data availability

The participant-level data used to generate the growth charts described here cannot be made publicly available as they contain protected health information from pediatric patients. The NF1-specific growth charts can be accessed at github.com/BGDlab, as well as the code used to produce them.

## Results

### Macrocephaly in NF1 Relative to CDC Charts and Community Control Cohort

The dataset contained 2180 observations from 305 individuals (167 male) with NF1, and 104,750 observations from 16,742 individuals (8809 male) in the community control cohort, all aged 0 to 3 years old (Table 1). Data were roughly evenly distributed across the age range (Supplementary Table 2).

The raw head circumference data for the NF1 and Well Child Visit cohorts are plotted on Figure 2A, along with CDC centile lines corresponding to the 3^rd^, 10^th^, 50^th^, 90^th^, and 97^th^ percentiles. While approximately 87% of the Well Child Visit head circumference measurements are contained between the 3^rd^ and 97^th^ centiles of the CDC growth charts, 23% of the NF1 data exceed the 97^th^ centile (Supplementary Table 3). Head circumference Z scores, as normed by the CDC charts, were consistently elevated in NF1 compared to the well child cohort across timepoints (*P*_adjusted_ < 0.05). The effect sizes varied nonlinearly with time, starting moderate at 1 month (*d* = 0.56), then small at four months (*d* = 0.28), moderate again at 15 months (*d* = 0.58), and finally large at 28 months (*d* = 0.8) (Figure 2B).

**Figure 2.**
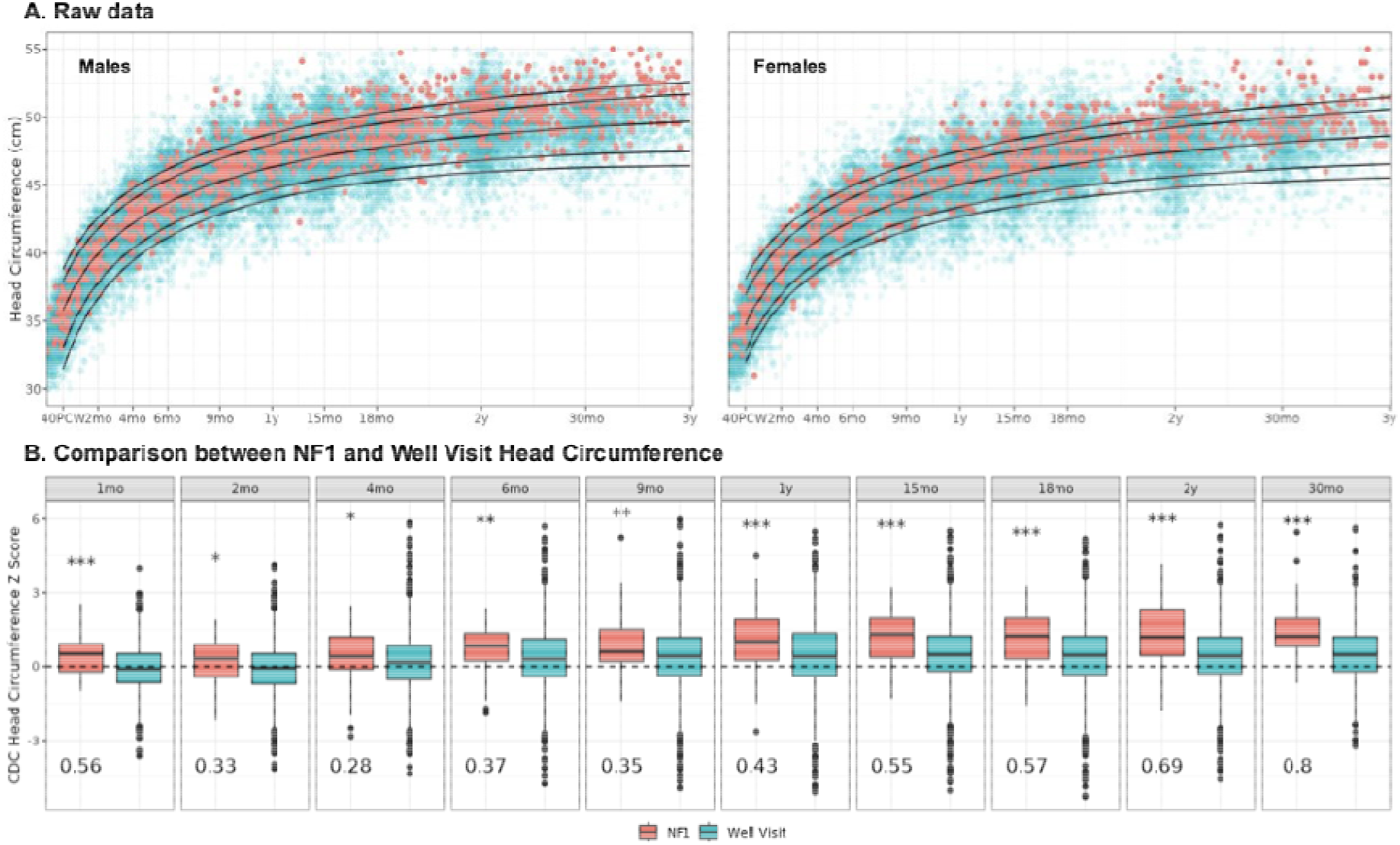
Head circumference data for NF1 and Well Visit Cohorts benchmarked using CDC charts. (A) Raw head circumference data plotted, colored by NF1 status, along with the 3^rd^, 10^th^, 50^th^, 90^th^, and 97^th^ centile lines from the CDC. (B) Boxplots comparing CDC head circumference Z score across timepoints for NF1 versus Well Visit. Cohen’s *d* displayed in the bottom left of each plot. *** = *P* < 0.001; ** = *P* < 0.01; * = *P* < 0.05, all adjusted for multiple comparisons via the Holm-Bonferroni method. Abbreviations: PCW = post-conceptional weeks

### NF1-Specific Growth Charts derived from LMSz

Using the recently described LMSz method,^26^ we produced NF1-specific charts for head circumference with the CDC head circumference charts as a reference. First, we iterated through a set of candidate models predicting CDC head circumference Z scores for the NF1 patients, selecting the one that minimized BIC (Supplementary Table 1; Figure 3A). The resulting model included only a linear term for age with no term for sex in the mu moment and only an intercept term for sigma. Using the LMS method, we back-transformed the centile lines for CDC head circumference Z score to head circumference measured in centimeters (Figure 3B). These centile lines contained the majority of the NF1 head circumference data (95.4% between the 3^rd^ and 97^th^ percentiles), unlike the original CDC centiles (76.7% between 3^rd^ and 97^th^ percentiles; Supplementary Table 3).

**Figure 3.**
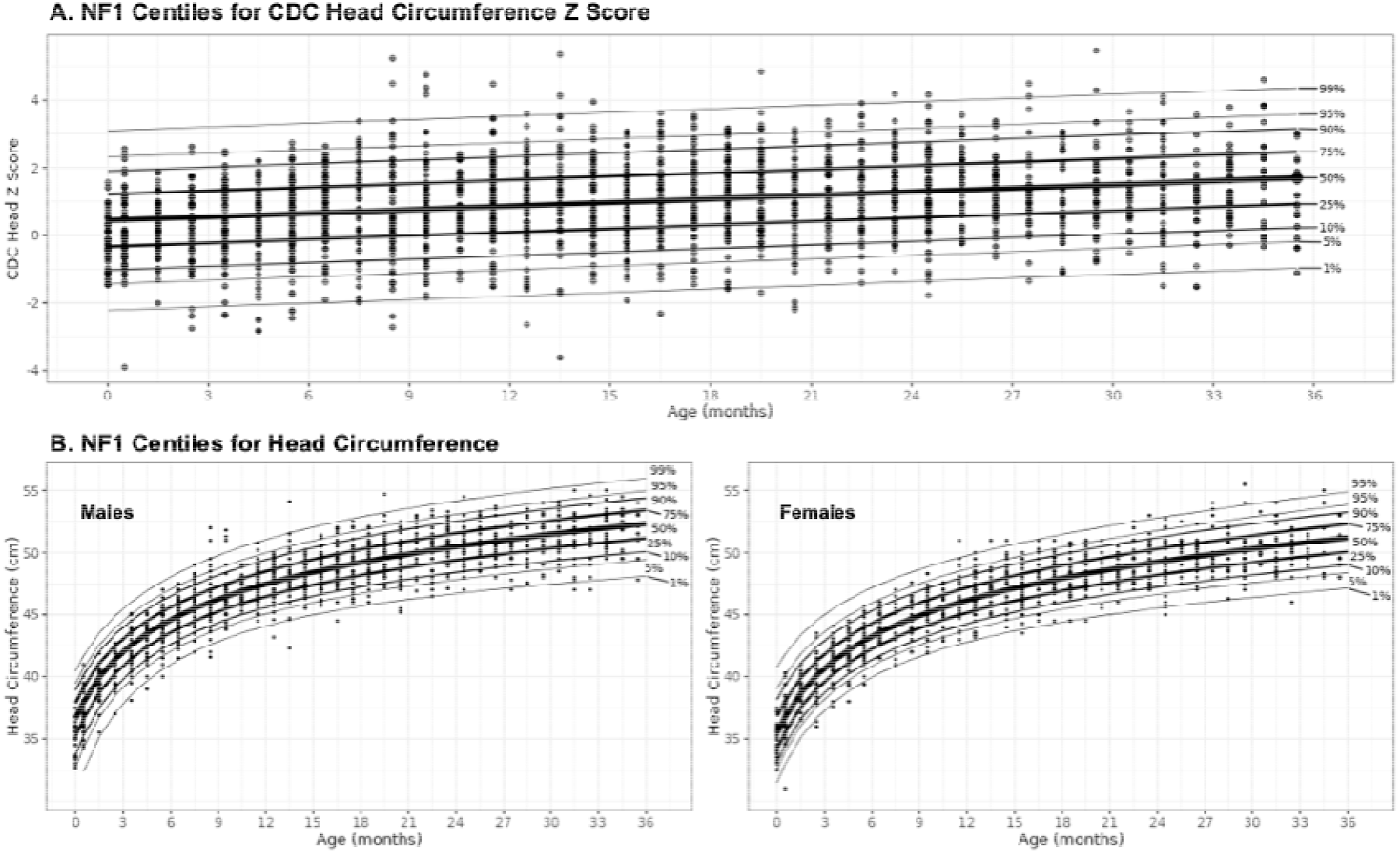
NF1-specific growth charts derived by applying the LMSz method to CDC head circumference charts. (A) NF1 centile lines for CDC head circumference Z score along with NF1 datapoints. (B) NF1 centile lines for head circumference measured in centimeters along with NF1 datapoints.

To compare between the NF1-specific growth charts and the CDC growth charts, we superimposed their plots (Figure 4A). By 35 months of age, the 10^th^ centile line for NF1 converges with the 50^th^ centile line in CDC, while the 50^th^ centile for NF1 exceeds the 90^th^ for CDC. By computing the derivative of the 50^th^ centile line, we examined the expected rate-of-growth for NF1 and CDC growth charts respectively (Figure 4B). While rate-of-growth appears similar over the first four months of life, the growth rate for NF1 decays at a slower pace compared to CDC and plateaus at a higher value.

**Figure 4.**
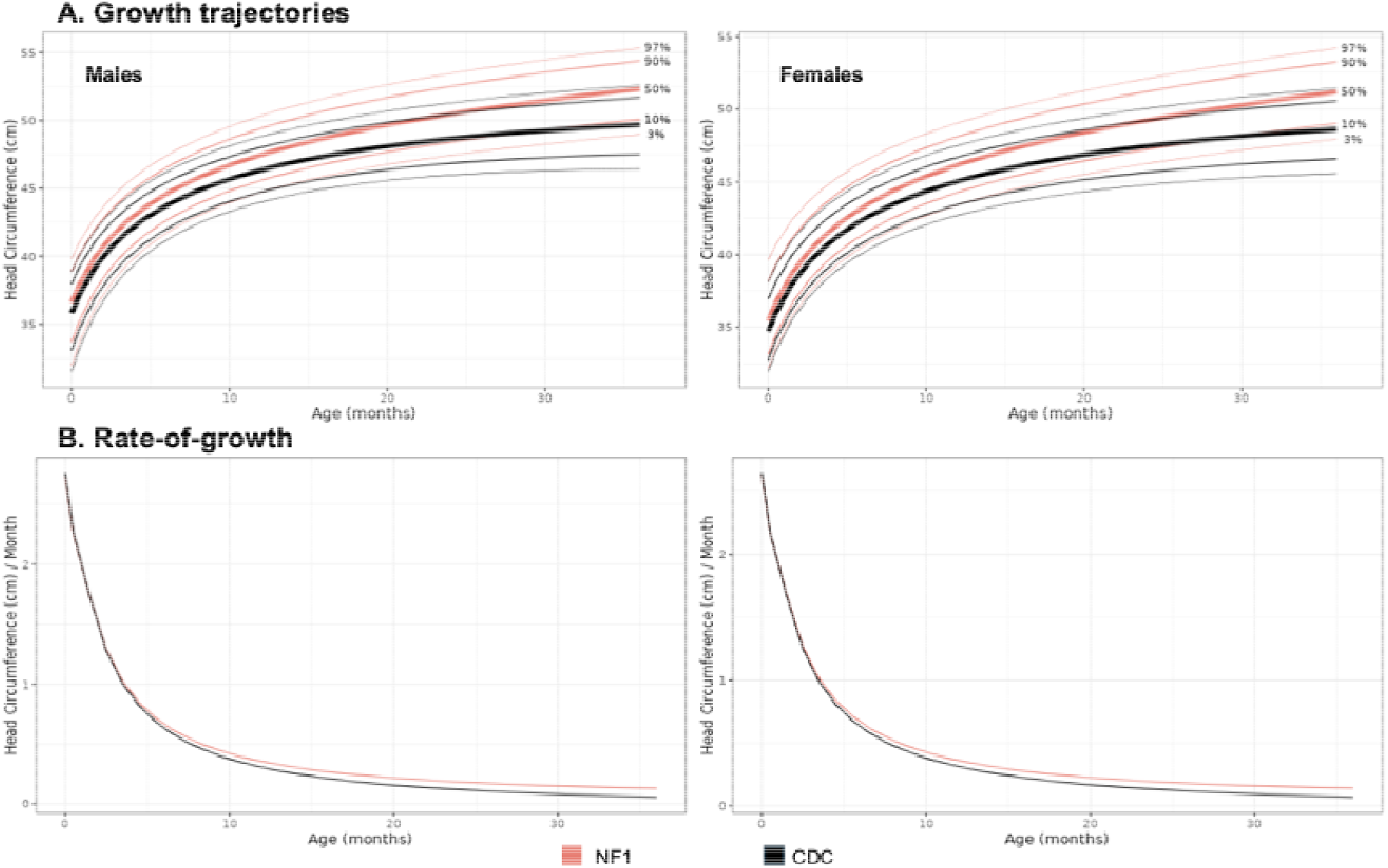
NF1 and CDC head circumference growth trajectories using LMSz method. (A) Centile lines corresponding to the 3^rd^, 10^th^, 50^th^, 90^th^, and 97^th^ percentiles respectively for NF1 and CDC. (B) Rate-of-growth for NF1 and CDC head circumference respectively.

### CHOP NF1 and Well Child Visit Growth Charts

As an alternative to using CDC growth charts, we constructed growth charts for the well child visit and NF1 head circumference data (Figure 5). In contrast to the CDC charts, age was represented in estimated post-conceptional days allowing for modeling of head circumference values in premature infants before 40 post-conceptional weeks (Figure 5A,B). Moreover, the NF1-specific charts were compared directly to a clinically-derived reference group. These charts demonstrate that head circumference for NF1 is slightly increased at the beginning of the lifespan, consistent with prior reports of an increased prevalence of macrocephaly at birth in NF1. Like the LMSz charts, median head circumference in NF1 begins to separate from the median clinical control trajectory starting around six months of age (Figure 5B).

**Figure 5.**
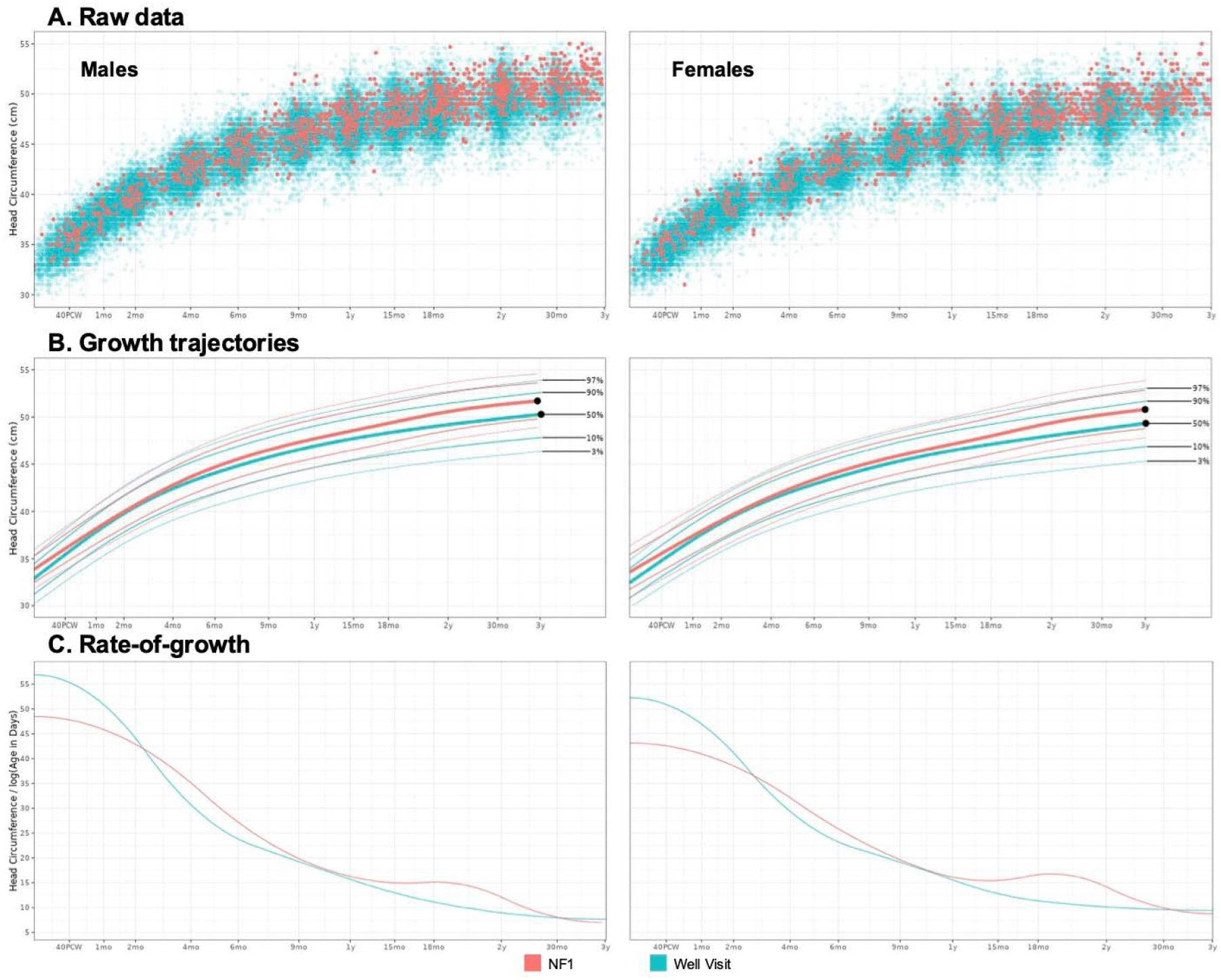
Head circumference growth trajectories for NF1 and community controls using CHOP growth charts. (A) Raw head circumference data plotted by post-conceptional age on a log scale. (B) Centile lines for NF1 and well child visit data. (C) Rate-of-growth for NF1 and Well Child Visit head circumference respectively.

Interestingly, and in contrast to the LMSz charts, rate-of-growth appears greater for community controls than NF1 patients for the first two months of life, before the curves intersect (Figure 5C). Following that intersection, there are two distinct time periods where NF1 head growth outpaces community control head growth for both males and females: from 2.5 to 9 months of age, and then 1 year to 30 months of age (Figure 5C). Despite slight differences in the shapes of the trajectories, NF1 head circumference Z scores computed using the LMSz and CHOP charts produced highly similar values (Supplementary Figure 8; intraclass correlation coefficient (ICC) = 0.97; *P* < 0.0001). Both charts also generalized well to an independent, Australian NF1 cohort (see Supplementary Materials).

### Microcephaly relative to NF1 Growth Expectations Relates to Developmental Delay

Finally, we aimed to see whether head circumference benchmarked against NF1 growth charts related to developmental delay in NF1 participants. Using 137 SWYC assessments obtained on the same day as head circumference measurements across 52 NF1 patients, we found that lower head circumference benchmarked upon LMSz NF1 growth charts related to developmental delay (Standardized beta = 0.24; *P* = 0.013) operationalized as low developmental quotient (Figure 6A). The same relationship was also seen when using head circumference Z scores from the CHOP charts instead of the LMSz charts (Standardized beta = 0.21; *P* = 0.028). This observation was in spite of the fact that none of these participants were microcephalic on the CDC growth charts (Figure 6B).

**Figure 6.**
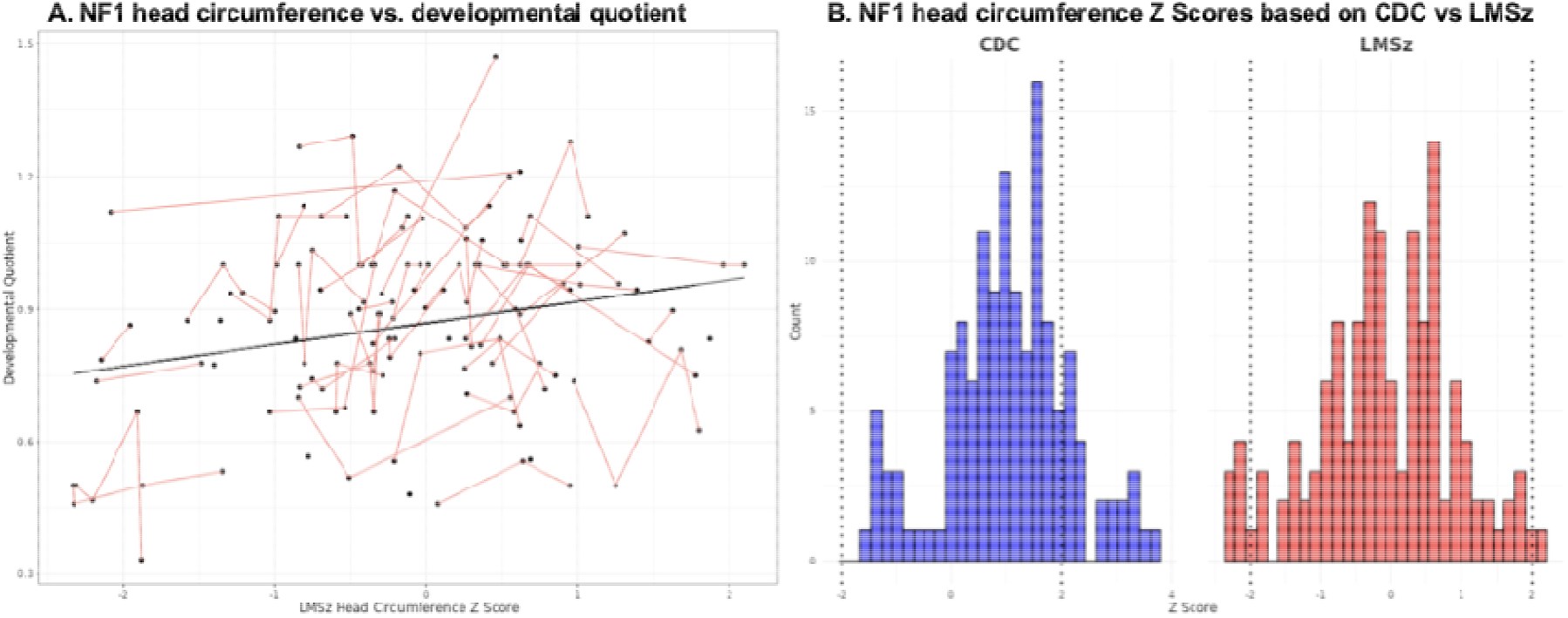
Relationship between NF1 head circumference benchmarked against NF1 growth expectations to developmental quotient. (A) Scatterplot plotting LMSz head circumference Z scores against developmental quotient (DQ) as derived from SWYC assessments. Each datapoint corresponds to an individual head circumference measurement aligned to a SWYC assessment conducted on the same day. Light red lines connect datapoints from the same individual. Black trendline obtained from linear mixed effects model fit to DQ with LMSz head circumference as a fixed effect and patient ID as a random effect. (B) Head circumference Z scores for the NF1 patients where SWYC assessment was also available computed using CDC and LMSz models respectively. Dotted lines correspond to commonly used definitions for microcephaly and macrocephaly, respectively.

## Discussion

Here, we present the first growth charts of NF1 head circumference that cover the first three years of life. We find that the difference in head growth in NF1 compared to typical development increases with time, and that there exist discrete periods of accelerated NF1 head growth over this period. Finally, we demonstrate that the construct of microcephaly relative to NF1-specific growth charts may be sensitive to developmental delay in NF1 patients, despite microcephaly relative to CDC charts being extremely uncommon in this population. These findings both clarify the timing of NF1 macrocephaly and suggest the potential clinical utility of NF1-specific growth charts as a more sensitive tool to detect abnormal patterns of growth in children with NF1.

### Timing of macrocephaly

While macrocephaly is among the most common clinical features in NF1,^8^ little is known about the timing of atypical head growth for these patients. Increased prevalence of macrocephaly has been observed at the time of birth in NF1,^32,33^ and our results similarly show significant differences in head circumference in the first months of life. However, we demonstrate that after four months of age, the head circumference differences become more exaggerated with time, both when comparing with the CDC growth charts and the community control cohort. The effect size comparing the differences between NF1 and community controls is at its lowest around four months (Cohen’s *d* = ∼0.2), but then steadily increases through 28 months (Cohen’s *d* = ∼0.8).

Since ASD is observed in NF1 in up to ∼30% of cases, it is tempting to speculate that brain overgrowth may be related to deficits in social cognition and motor delays also seen in NF1.^18,34^ Early brain overgrowth is also a hypothesized feature of ASD, though more recent evidence implies that macrocephaly is only seen in a subset of ASD cases sometimes associated with specific genetic syndromes.^35,36^ NF1 is an especially compelling model to investigate biological underpinnings of ASD compared to other genetic disorders associated with ASD (e.g. fragile X, 22q11.2 deletion syndrome) because intelligence is relatively preserved in NF1.^37,38^ Some neuroimaging studies have linked elevated brain structural volumes in NF1 with social deficits, though these studies were in relatively small samples and were limited to adolescent/adult populations.^18,39^ While outside the scope of the present study, a compelling future direction is to better understand the neurocognitive associations of brain growth abnormalities in NF1, both to inform targeted early interventions for NF1 patients as well as to elucidate biological mechanisms of developmental psychopathology more generally.

### Rate-of-growth

We present two complementary methods for deriving NF1 head circumference growth charts using the recently described LMSz method and an approach based on methods used by the WHO, respectively. The LMSz method borrows information about how children typically grow from another established growth chart (i.e., CDC charts) and updates the growth curves using data from patients with a specific genetic syndrome (i.e., NF1). A key limitation of these charts is incomplete modeling of head circumference before 40 post-conceptional weeks (as the CDC charts start at birth) as well as the potential for bias in any contemporary clinical population relative to CDC norms.^15^ Several studies have demonstrated that the CDC charts underestimate centiles even among typically developing children, complicating some of the existing literature on early brain overgrowth in ASD.^15,28^

To address some of these limitations, we also present CHOP growth charts modelling head circumference as a function of post-conceptional age. Interestingly, this approach suggests that head growth is actually slower in the first two months of life in NF1 compared to the control group. A possible explanation is that accelerated head growth is occurring earlier in gestation in NF1, considering that NF1 children often present with macrosomia at birth.^33^ Both modelling approaches highlight sustained growth in NF1, as the NF1 growth curves decelerate at a slower pace compared to controls. However, while the LMSz approach models NF1-specific growth differences with only a linear term for age, the CHOP charts suggests greater nonlinearity, with distinct time periods of accelerated growth relative to controls in NF1, from 2.5 to 9 months of age, and then 1 year to 30 months of age. Further research in independent samples is required to verify the generalizability of these findings. Both methods produce very similar Z scores in practice.

### Clinical implications

In current clinical practice, common growth references like the CDC charts are applied to all patients regardless of genotype, with very few exceptions.^40^ In theory, this practice could decrease sensitivity for early signs of abnormal growth if a patient has a genetic syndrome with differing growth expectations from typical development.^20^ We provide evidence for decreased sensitivity of the CDC charts in detecting abnormal head growth in children with NF1.

We observed a relationship between head circumference relative to NF1 growth expectations and developmental delay, even though none of the participants with developmental delay had microcephaly as defined by the CDC charts. NF1 participants with small head circumference, relative to their cohort, may be missed on CDC charts that compare their growth to typically developing children, neglecting the fact that the genetic syndrome results in an expectation of more rapid head growth. Adoption of NF1-specific growth charts could facilitate early identification and intervention for NF1 children who are not growing as expected.^26^

Some limitations of the current study should be noted. First, the data used to produce NF1 head circumference growth charts were collected from patients seen at a multidisciplinary NF Program in an urban, tertiary academic medical center, which likely introduces some selection bias, as many of the patients may have been referred for management. For example, these participants may be less representative of NF1 patients with milder, subclinical symptoms who often go undiagnosed. Reassuringly, the NF1 growth charts generalized to an independent NF1 cohort recruited in a neurogenetics clinic in Australia. Second, while our data contain repeated head circumference measurements from the same participants, we do not model the dependence between these repeated measurements when constructing growth charts. While this approach is taken in many other descriptive growth studies and does not bias resulting growth charts,^26,28,29,40,41^ explicit modeling of longitudinal data could help clarify the expected trajectories of individual patients. Like other growth charts that do not model longitudinal effects, longitudinal monitoring of centiles (e.g. evaluation of “centile crossing”) should be approached with caution as phenomena like regression to the mean can complicate the interpretation.^42^ Third, the developmental screen data is limited in its reliance on subjective reports from parents on milestone completion and also in sample size. Finally, while macrocephaly correlates with megalencephaly,^9,10,43^ it is not possible to directly infer insights into brain development from a study of head circumference.

In sum, we provide the first publicly available growth charts of head circumference for NF1 that cover the first three years of life and demonstrate their increased sensitivity to abnormal growth patterns in youth with NF1. Future work should clarify how head growth relates to cognitive and social skills development in NF1. Another important direction for future work is to directly investigate the brain correlates of interindividual differences in head circumference. No quantitative neuroimaging study to our knowledge has examined brain development over the first three years of life in NF1. NF1-specific brain growth charts, potentially leveraging clinical neuroimaging,^23,44^ would provide valuable insight into the neuropathology of NF1.

## Acknowledgements

This work was funded by a US Department of Defense Neurofibromatosis Synergistic Idea Award (PIs Garg, Payne, Fisher), NIMH R01MH134896 (PI Alexander-Bloch), and the CHOP Research Institute.

