## Supplementary Materials for "NF1-specific growth charts for head circumference over the first three years of life"

Ayan S. Mandal MD PhD^1,2,3^, Russell T. Shinohara PhD^4^, Benjamin Jung PhD^2,3^, Margaret Gardner^2,3^, Habib E. Akouri^1^, Benjamin E. Yerys PhD^2,5,6^, Whitney Guthrie PhD^5^, Kelly M. Janke PhD, ABPP-CN^2^, John D. Herrington PhD^2,5^, Matthew C. Hocking PhD^1,2^, Gareth Ball PhD^7,8^, Jonathan M. Payne PsyD^7,8^, Kathryn N. North MD^7,8^, Nils Muhlert PhD^9^, Shruti Garg PhD^10^, Jakob Seidlitz PhD^2,3,11^, Michael J. Fisher MD^1,12^, Aaron Alexander-Bloch MD PhD^2,3,11^

1 Perelman School of Medicine, University of Pennsylvania, Philadelphia, PA, USA

2 Department of Child and Adolescent Psychiatry and Behavioral Science, The Children’s Hospital of Philadelphia, Philadelphia, PA, USA.

3 Lifespan Brain Institute, Children’s Hospital of Philadelphia and Penn Medicine, Philadelphia, PA, USA

4 Penn Statistics in Imaging and Visualization Center, Department of Biostatistics, Epidemiology, and Informatics, University of Pennsylvania, Philadelphia, PA, USA

5 Center for Autism Research, The Children’s Hospital of Philadelphia and Penn Medicine, Philadelphia PA, USA

6 Advancing Transition and Learning for Adult Success Center, The Children’s Hospital of Philadelphia and Penn Medicine, Philadelphia, PA, USA

7 Developmental Imaging, Murdoch Children’s Research Institute, Melbourne, Victoria, Australia

8 Department of Paediatrics, University of Melbourne, Melbourne, Victoria, Australia

9 Division of Psychology, Communication & Human Neurosciences, Faculty of Biology, Medicine and Health, University of Manchester, UK

10 Division of Psychology and Mental Health, Faculty of Biology, Medicine and Health, University of Manchester and Manchester University Foundation NHS Trust, Manchester, UK

11 Department of Psychiatry, University of Pennsylvania, Philadelphia, PA, USA

12 Division of Oncology, Children’s Hospital of Philadelphia, Philadelphia, PA, USA

**Supplementary Methods and Results**

*Validation in independent cohort*

     To test the generalizability of the NF1 growth charts, we plotted head circumference data from NF1 participants seen at a neurogenetics clinic in Australia onto the charts. Participant recruitment, selection, and exclusion criteria are described in more detail in a prior publication.^1^ Briefly, children who met the NIH diagnostic criteria for NF1 were prospectively recruited at the Neurogenetics Clinic at The Children’s Hospital at Westmead for a longitudinal study involving regular neurodevelopmental assessments, including head circumference measurements, until up to 7 years of age. Children with inadequate English skills, hearing problems, and/or optic gliomas were excluded. We used data from participants under the age of 3.

     The Australian NF1 dataset included 209 head circumference measurements from 90 participants (51 males). Measurements were obtained at specific age intervals, including 5-6 months, 9-11 months, 14-16 months, 20-22 months, and 29-31 months. Age was recorded in months and post-conceptional age was computed in the same way as for the Well Child Visit and NF1 Cohorts from CHOP. Gestational age at birth was assumed to be 40 weeks as this was unavailable for all participants. Centiles were computed for each head circumference observation using the LMSz and CHOP NF1 growth charts. We also applied asymptotic one-sample Kolmogorov-Smirnov tests to determine whether the centiles fit a uniform distribution, using the   first datapoint for each participant to eliminate repeated (dependent) measures.

     Both the LMSz and CHOP NF1 growth charts accurately characterized growth trajectories in the Australian NF1 cohort (Supplementary Figure 9). 95.6% and 94.5% of participants were between the 3rd and 97th percentiles for the LMSz and CHOP growth charts, respectively (Supplementary Table 4). Centiles computed from each approach appeared to fit a uniform distribution (for LMSz: *D* = 0.078; *P* = 0.65; for CHOP: *D* = 0.092; *P* = 0.42)

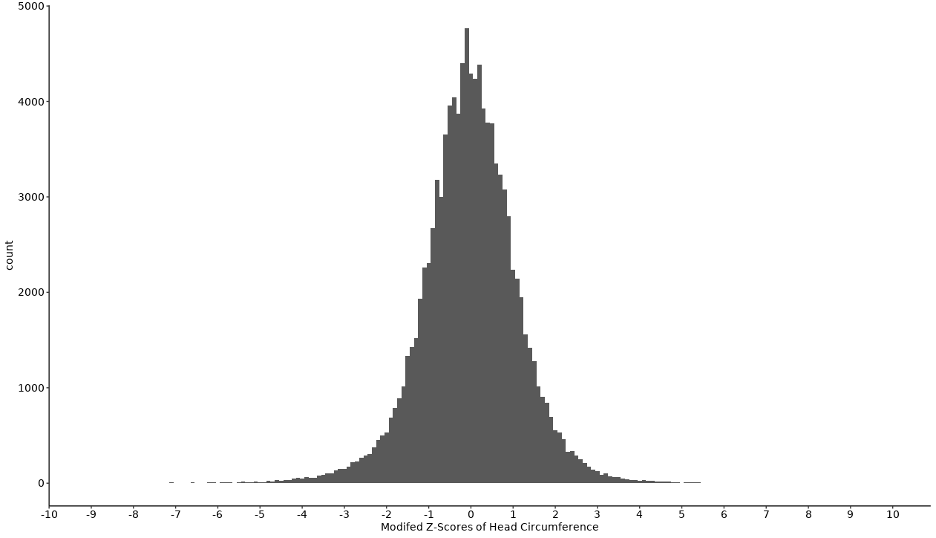

**Supplementary Figure 1. Modified Z-Scores of Head Circumference.** X-axis was limited to show values between -10 and 10. Based on this histogram, values lower than -5 and above 5 were excluded.

 
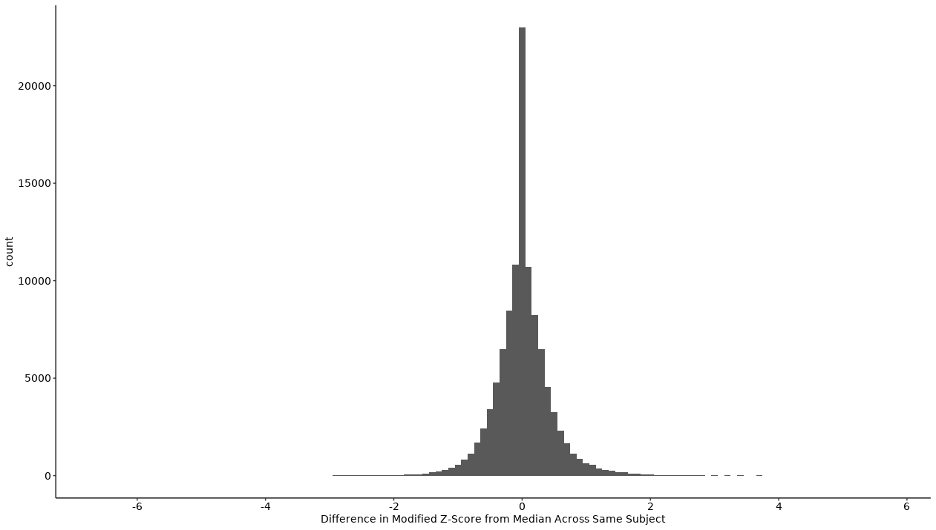

**Supplementary Figure 2. Difference in Modified Z-Score from Median Across Same Subject.** Based on this histogram, values lower than -2 and above 2 were excluded.

 
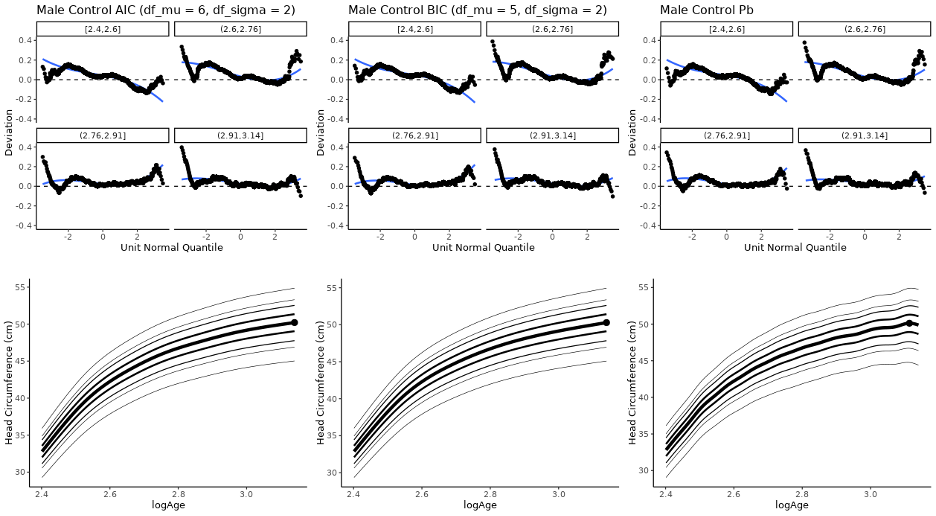

**Supplementary Figure 3. Worm plots and growth charts of GAMLSS models of male well-visit data optimizing AIC, BIC, and penalized b-spline respectively.** Of these, the model optimizing BIC was selected on account of similar worm plots to the other two, smoother curves compared to penalized b-spline, and simpler model compared to the one optimizing AIC. Confidence bands are not included in the worm plots as the data contain repeated measurements which complicates their interpretation.

 
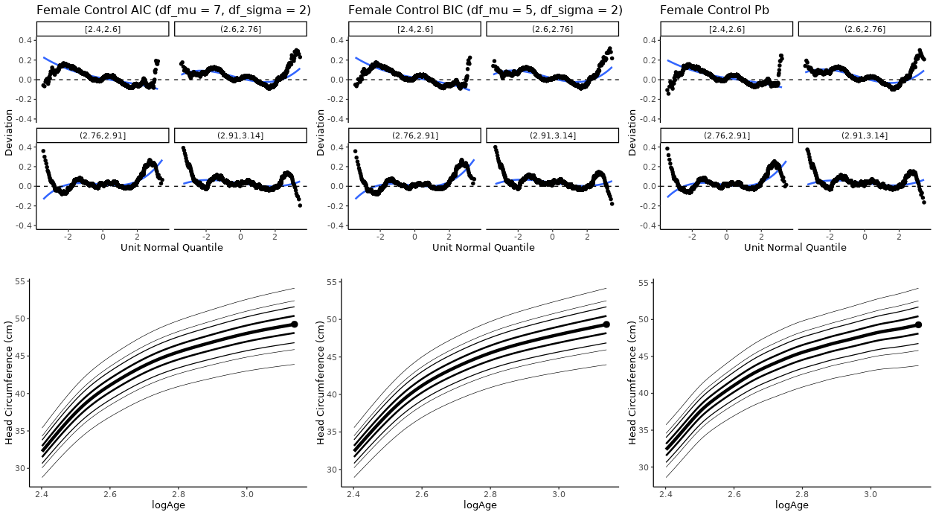

**Supplementary Figure 4. Worm plots and growth charts of GAMLSS models of female well-visit data optimizing AIC, BIC, and penalized b-spline respectively.** Of these, the model optimizing BIC was selected on account of similar worm plots to the other two, smoother curves compared to penalized b-spline, and simpler model compared to the one optimizing AIC.

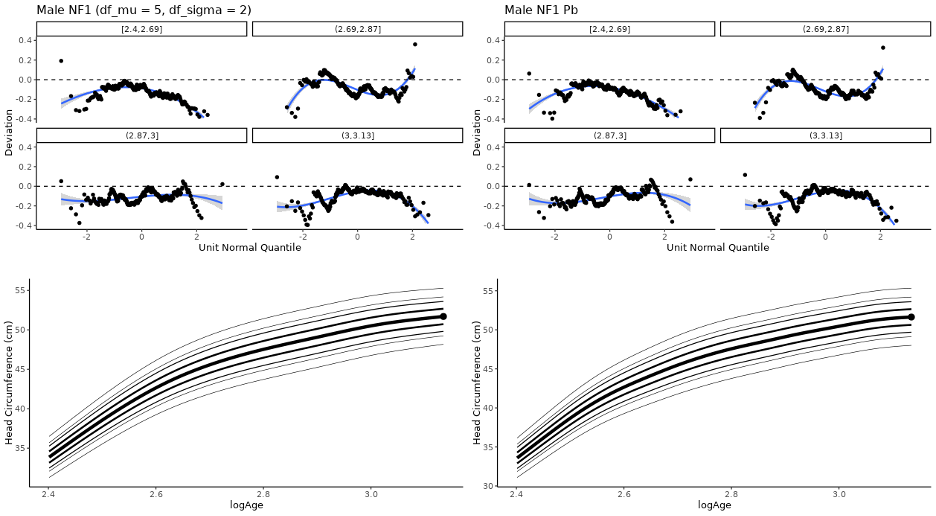

**Supplementary Figure 5. Worm plots and growth charts of GAMLSS models of male NF1 data optimizing AIC/BIC and penalized b-spline respectively.** The same model was selected by AIC and BIC. On account of smoother growth curves and a simpler model, the natural spline model optimizing AIC and BIC was chosen.

 
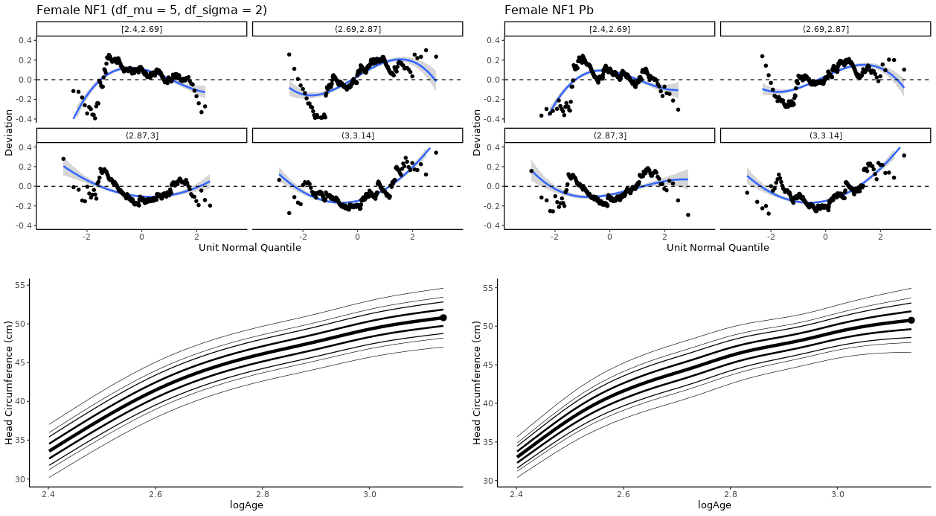

**Supplementary Figure 6. Worm plots and growth charts of GAMLSS models of female NF1 data optimizing AIC/BIC and penalized b-spline respectively.** The same model was selected by AIC and BIC. On account of smoother growth curves and a simpler model, the natural spline model optimizing AIC and BIC was chosen.

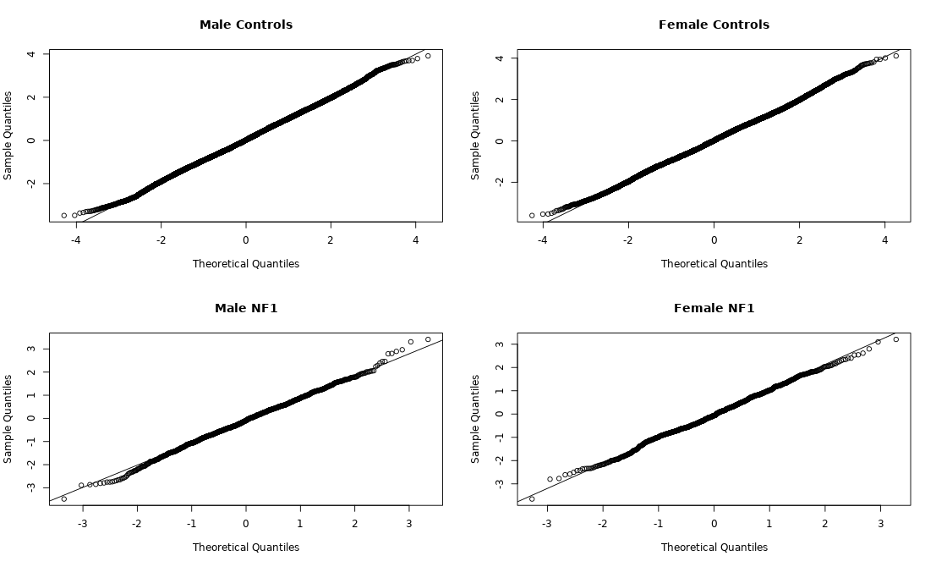

**Supplementary Figure 7. Q-Q plots for selected GAMLSS models for each cohort.**

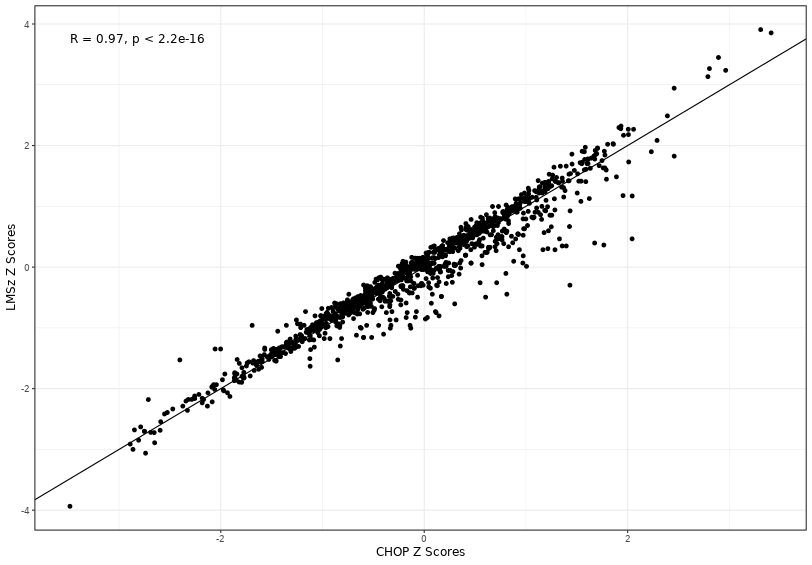

**Supplementary Figure 8. Scatterplot of Z scores for the NF1 head circumference measurements computed using the LMSz and CHOP charts respectively.** Black line reflects the relationship if these scores were equal at every observation. An intraclass correlation coefficient (ICC) computed on these data confirmed a strong relationship (ICC2 = 0.97; *P* < 0.0001).

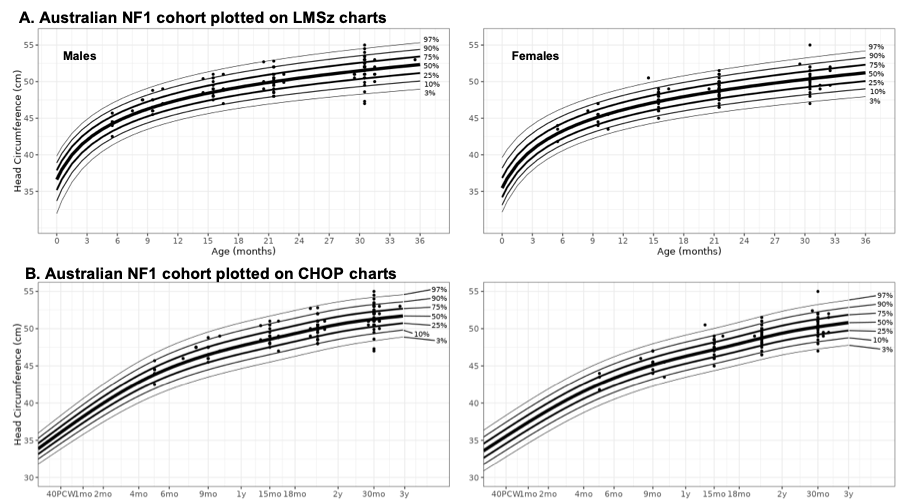
**Supplementary Figure 9. Validation of NF1 growth charts using an independent, Australian NF1 cohort.**

| **Mu formulas** | **Sigma formulas** | **AIC** | **BIC** |
| --- | --- | --- | --- |
| ~age + sex + age*sex | ~age + sex | 927.7605 | 967.5989 |
| ~age + sex | ~age + sex | 925.761 | 959.9082 |
| ~age | ~age + sex | 923.8091 | 952.2651 |
| ~sex | ~age + sex | 947.1055 | 975.5615 |
| ~1 | ~age + sex | 945.1595 | 967.9243 |
| ~0 | ~age + sex | 1102.2507 | 1119.3243 |
| ~pb(age) + sex + age*sex | ~age + sex | 928.5737 | 986.8731 |
| ~pb(age) + sex | ~age + sex | 926.5729 | 979.1646 |
| ~pb(age) | ~age + sex | 924.5991 | 971.3577 |
| ~ns(age,2) + sex + age*sex | ~age + sex | 925.1406 | 970.6702 |
| ~ns(age,2) + sex | ~age + sex | 923.1435 | 962.9819 |
| ~ns(age,2) | ~age + sex | 921.1931 | 955.3403 |
| ~ns(age,3) + sex + age*sex | ~age + sex | 926.897 | 978.1178 |
| ~ns(age,3) + sex | ~age + sex | 924.8985 | 970.4281 |
| ~ns(age,3) | ~age + sex | 922.9483 | 962.7867 |
| ~ns(age,4) + sex + age*sex | ~age + sex | 928.4282 | 985.3402 |
| ~ns(age,4) + sex | ~age + sex | 926.4306 | 977.6514 |
| ~ns(age,4) | ~age + sex | 924.4951 | 970.0247 |
| ~ns(age,5) + sex + age*sex | ~age + sex | 930.0798 | 992.683 |
| ~ns(age,5) + sex | ~age + sex | 928.0831 | 984.9951 |
| ~ns(age,5) | ~age + sex | 926.1528 | 977.3736 |
| ~age + sex + age*sex | ~age | 925.7896 | 959.9368 |
| ~age + sex | ~age | 923.7901 | 952.2461 |
| ~age | ~age | 921.8381 | 944.6029 |
| ~sex | ~age | 945.1142 | 967.879 |
| ~1 | ~age | 943.1681 | 960.2417 |
| ~0 | ~age | 1100.2514 | 1111.6338 |
| ~pb(age) + sex + age*sex | ~age | 926.5834 | 979.1832 |
| ~pb(age) + sex | ~age | 924.5827 | 971.4749 |
| ~pb(age) | ~age | 922.6089 | 963.6686 |
| ~ns(age,2) + sex + age*sex | ~age | 923.1516 | 962.99 |
| ~ns(age,2) + sex | ~age | 921.1544 | 955.3016 |
| ~ns(age,2) | ~age | 919.204 | 947.66 |
| ~ns(age,3) + sex + age*sex | ~age | 924.9061 | 970.4357 |
| ~ns(age,3) + sex | ~age | 922.9076 | 962.746 |
| ~ns(age,3) | ~age | 920.9573 | 955.1045 |
| ~ns(age,4) + sex + age*sex | ~age | 926.4366 | 977.6574 |
| ~ns(age,4) + sex | ~age | 924.4391 | 969.9687 |
| ~ns(age,4) | ~age | 922.5035 | 962.3419 |
| ~ns(age,5) + sex + age*sex | ~age | 928.0936 | 985.0056 |
| ~ns(age,5) + sex | ~age | 926.0968 | 977.3176 |
| ~ns(age,5) | ~age | 924.1664 | 969.696 |
| ~age + sex + age*sex | ~sex | 925.7612 | 959.9084 |
| ~age + sex | ~sex | 923.7616 | 952.2176 |
| ~age | ~sex | 921.8097 | 944.5745 |
| ~sex | ~sex | 945.8416 | 968.6064 |
| ~1 | ~sex | 943.8935 | 960.9671 |
| ~0 | ~sex | 1109.9077 | 1121.2901 |
| ~pb(age) + sex + age*sex | ~sex | 926.5733 | 979.1798 |
| ~pb(age) + sex | ~sex | 924.5728 | 971.4726 |
| ~pb(age) | ~sex | 922.5992 | 963.6666 |
| ~ns(age,2) + sex + age*sex | ~sex | 923.1407 | 962.9791 |
| ~ns(age,2) + sex | ~sex | 921.1436 | 955.2908 |
| ~ns(age,2) | ~sex | 919.1932 | 947.6492 |
| ~ns(age,3) + sex + age*sex | ~sex | 924.898 | 970.4276 |
| ~ns(age,3) + sex | ~sex | 922.8996 | 962.738 |
| ~ns(age,3) | ~sex | 920.9494 | 955.0966 |
| ~ns(age,4) + sex + age*sex | ~sex | 926.4282 | 977.649 |
| ~ns(age,4) + sex | ~sex | 924.4307 | 969.9603 |
| ~ns(age,4) | ~sex | 922.4952 | 962.3336 |
| ~ns(age,5) + sex + age*sex | ~sex | 928.0798 | 984.9918 |
| ~ns(age,5) + sex | ~sex | 926.0831 | 977.3039 |
| ~ns(age,5) | ~sex | 924.1528 | 969.6824 |
| ~age + sex + age*sex | ~1 | 923.7908 | 952.2468 |
| ~age + sex | ~1 | 921.7913 | 944.5561 |
| **~age** | **~1** | **919.8392** | **936.9128** |
| ~sex | ~1 | 943.8629 | 960.9365 |
| ~1 | ~1 | 941.9146 | 953.297 |
| ~0 | ~1 | 1107.9137 | 1113.6049 |
| ~pb(age) + sex + age*sex | ~1 | 924.5825 | 971.4872 |
| ~pb(age) + sex | ~1 | 922.582 | 963.7802 |
| ~pb(age) | ~1 | 920.6084 | 955.9749 |
| ~ns(age,2) + sex + age*sex | ~1 | 921.1516 | 955.2988 |
| ~ns(age,2) + sex | ~1 | 919.1544 | 947.6104 |
| **~ns(age,2)** | **~1** | **917.204** | **939.9688** |
| ~ns(age,3) + sex + age*sex | ~1 | 922.9068 | 962.7452 |
| ~ns(age,3) + sex | ~1 | 920.9084 | 955.0556 |
| ~ns(age,3) | ~1 | 918.9582 | 947.4142 |
| ~ns(age,4) + sex + age*sex | ~1 | 924.4366 | 969.9662 |
| ~ns(age,4) + sex | ~1 | 922.4391 | 962.2775 |
| ~ns(age,4) | ~1 | 920.5035 | 954.6507 |
| ~ns(age,5) + sex + age*sex | ~1 | 926.0936 | 977.3144 |
| ~ns(age,5) + sex | ~1 | 924.0968 | 969.6264 |
| ~ns(age,5) | ~1 | 922.1664 | 962.0048 |
| ~age + sex + age*sex | ~0 | 934.3267 | 957.0915 |
| ~age + sex | ~0 | 932.3273 | 949.4009 |
| ~age | ~0 | 930.3906 | 941.773 |
| ~sex | ~0 | 963.4829 | 974.8653 |
| ~1 | ~0 | 961.5571 | 967.2483 |
| ~0 | ~0 | 1285.0129 | 1285.0129 |
| ~pb(age) + sex + age*sex | ~0 | 933.3605 | 974.574 |
| ~pb(age) + sex | ~0 | 931.3615 | 966.8684 |
| ~pb(age) | ~0 | 929.4104 | 959.0856 |
| ~ns(age,2) + sex + age*sex | ~0 | 930.2437 | 958.6997 |
| ~ns(age,2) + sex | ~0 | 928.2474 | 951.0122 |
| ~ns(age,2) | ~0 | 926.3119 | 943.3855 |
| ~ns(age,3) + sex + age*sex | ~0 | 931.9254 | 966.0726 |
| ~ns(age,3) + sex | ~0 | 929.9275 | 958.3835 |
| ~ns(age,3) | ~0 | 927.9922 | 950.757 |
| ~ns(age,4) + sex + age*sex | ~0 | 933.3148 | 973.1532 |
| ~ns(age,4) + sex | ~0 | 931.318 | 965.4652 |
| ~ns(age,4) | ~0 | 929.4016 | 957.8576 |
| ~ns(age,5) + sex + age*sex | ~0 | 934.8699 | 980.3995 |
| ~ns(age,5) + sex | ~0 | 932.8741 | 972.7125 |
| ~ns(age,5) | ~0 | 930.9642 | 965.1114 |

**Supplementary Table 1. AIC and BIC of candidate models for LMSz method.** Models that minimized AIC and BIC respectively are bolded. Degrees of freedom for the natural spline models are indicated in the parentheses. Abbreviations: pb = penalized b-spline; ns = natural spline.

| **Cohort** | **< 1mo** | **1- 2 mo** | **2 – 4 mo** | **4 – 6 mo** | **6 – 9 mo** | **9 mo – 1y** | **1 y – 15 mo** | **15 – 18 mo** | **18 mo – 2y** | **2y – 30 mo** | **30 mo – 3y** |
| --- | --- | --- | --- | --- | --- | --- | --- | --- | --- | --- | --- |
| NF1 | 131 | 63 | 120 | 153 | 194 | 221 | 200 | 192 | 404 | 288 | 214 |
| Well Visit | 10644 | 7155 | 9730 | 9923 | 10972 | 11192 | 10711 | 10209 | 12063 | 8962 | 3205 |

**Supplementary Table 2. Number of head circumference measurements within various age bins for NF1 and Well Visit cohorts.**

| **Growth Chart** | **Cohort** | **3^rd^ Percentile** | **10^th^ Percentile** | **50^th^ Percentile** | **90^th^ Percentile** | **97^th^ Percentile** |
| --- | --- | --- | --- | --- | --- | --- |
| CDC | Well Visit | 4.3% | 8.9% | 40.6% | 82.5% | 91.7% |
| CDC | NF1 | 0.3% | 2.6% | 13.1% | 56.7% | 77% |
| LMSz | NF1 | 2.6% | 7.5% | 49.2% | 90.2% | 98% |
| CHOP | NF1 | 2.6% | 8.2% | 49.2% | 91.1% | 98.4% |

**Supplementary Table 3. Percentage of participants under the 3^rd^, 10^th^, 50^th^, 90^th^, and 97^th^ percentiles for head circumference across growth chart approaches.** The percentage of participants above these percentiles can be computed by subtracting these numbers from 100% (e.g. 23% of NF1 participants were above the 97^th^ percentile on the CDC charts). Centiles using each approach were constructed for each observation and the median was computed across observations for each participant.

| **Growth Chart** | **3^rd^ Percentile** | **10^th^ Percentile** | **50^th^ Percentile** | **90^th^ Percentile** | **97^th^ Percentile** |
| --- | --- | --- | --- | --- | --- |
| CDC | 0% | 1.1% | 12.2% | 46.7% | 73.3% |
| LMSz | 2.2% | 7.8% | 43.3% | 92.2% | 97.8% |
| CHOP | 2.2% | 7.8% | 43.3% | 92.2% | 96.7% |

**Supplementary Table 4. Percentage of Australian NF1 cohort under the 3^rd^, 10^th^, 50^th^, 90^th^, and 97^th^ percentiles for head circumference across growth chart approaches.** The percentage of participants above these percentiles can be computed by subtracting these numbers from 100%. Centiles using each approach were constructed for each observation and the median was computed across observations for each participant.
